# Variants in *PRKAR1B* cause a neurodevelopmental disorder with autism spectrum disorder,apraxia, and insensitivity to pain

**DOI:** 10.1101/2020.09.10.20190314

**Authors:** Felix Marbach, Georgi Stoyanov, Florian Erger, Jill A. Rosenfeld, Erin Torti, Chad Haldeman-Englert, Evgenia Sklirou, Elena Kessler, Sophia Ceulemans, Stanley F. Nelson, Julian A. Martinez-Agosto, Christina G.S. Palmer, Rebecca H. Signer, Undiagnosed Diseases Network, Marisa V. Andrews, Dorothy K. Grange, Rebecca Willaert, Richard Person, Aida Telegrafi, Olivier Lichtarge, Panagiotis Katsonis, Amber Stocco, Christian P. Schaaf

**Affiliations:** Institute of Human Genetics, Heidelberg University, Heidelberg, Germany; Faculty of Medicine, University of Cologne, Cologne, Germany; Institute of Human Genetics, University of Cologne, University Hospital Cologne, Cologne, Germany; Department of Molecular and Human Genetics, Baylor College of Medicine, Houston, Texas, USA; Baylor Genetics Laboratory, Houston, Texas, USA; GeneDX, Gaithersburg, Maryland, USA; Mission Fullerton Genetics Center, Asheville, North Carolina, USA; Department of Pediatrics, University of Pittsburgh School of Medicine, Pittsburgh, Pennsylvania, USA; Genetics / Dysmorphology, Rady Children’s Hospital, San Diego, California, USA; Department of Human Genetics, David Geffen School of Medicine at UCLA, Los Angeles, California, USA; Department of Psychiatry & Biobehavioral Sciences, David Geffen School of Medicine at UCLA, Los Angeles, California, USA; Institute for Society and Genetics, UCLA, Los Angeles, California, USA; Division of Genetics and Genomic Medicine, Department of Pediatrics, Washington University School of Medicine, Saint Louis, Missouri, USA; INTEGRIS Pediatric Neurology, Oklahoma City, Oklahoma, USA

## Abstract

**Purpose:** We characterize the phenotypes of six unrelated individuals with intellectual disability and autism spectrum disorder, who carry heterozygous missense-variants of the *PRKAR1B* gene.

**Methods:** Variants of *PRKAR1B* were identified by single-exome or trio-exome analysis. We contacted the families and physicians of the six individuals in order to collect clinical and phenotypic information.

**Results:** *PRKAR1B* encodes the R1β subunit of the cyclic AMP-dependent protein kinase A (PKA), and is predominantly expressed in the central nervous system. Recent studies of patient cohorts with neurodevelopmental disorders found significant enrichment of *de novo* missense variants in *PRKAR1B*, and *in vivo* studies of the murine ortholog demonstrated altered hippocampal function and reduced neurogenic inflammation and long-term nociceptive pain in R1β-deficient mice.

In our study, *de novo* origin of the *PRKAR1B*-variants could be confirmed in five out of six individuals, and four carried the same heterozygous *de novo* variant c.1003C>T (p. Arg335Trp; NM_001164760). Global developmental delay, autism spectrum disorder, and apraxia/dyspraxia has been reported in all six, and reduced pain sensitivity was found in three individuals carrying the c.1003C>T variant.

**Conclusion:** Our study provides strong evidence for a novel, *PRKAR1B*-related neurodevelopmental disorder.

## Introduction

The gene *PRKAR1B* (Protein Kinase cAMP-Dependent Type I Regulatory Subunit Beta) encodes a regulatory subunit of the cyclic AMP-dependent protein kinase A protein complex (PKA), which is a nearly universal cellular component in eukaryotes^1^. PKA is a heterotetramer of two regulatory (R) and two catalytic (C) subunits, which, upon activation of PKA by cAMP, phosphorylates serine or threonine residues of different target proteins. Four R subunits (RIα, RIβ, RIIα and RIIβ) and six principal C subunits (Cα1, Cα2, Cβ1–4) are expressed in humans, and cell type-specific expression of different subunits changes the composition and thereby intracellular localization and substrate specificity of PKA isoforms^2^. R subunits serve as cAMP receptors and facilitate the spatial localization of PKA within the cell by binding different A-Kinase anchoring proteins (AKAPs)^3^. The subunit RIβ is primarily expressed in the brain^4,5^, with the highest levels of expression in the cerebral cortex and hypothalamus^6^.

The first functional study of R1β involving R1β-deficient mice dates back 25 years, with mice deficient of the murine ortholog of R1β showing altered hippocampal long-term depression and depotentiation^7^. Downregulation of R1β in murine hippocampal cultures was found to reduce the phosphorylation of CREB^8^, a transcription factor implicated in long-term memory formation^9^. Furthermore, R1β-deficient mice showed diminished nociceptive pain and inflammation in the setting of persistent tissue injury, although the reaction to acute nociceptive stimuli was unaffected^10^.

A missense variant in *PRKAR1B* has been associated with a rare hereditary neurodegenerative disorder in humans with R1β-positive inclusions in affected neurons^11,12^, but there is also mounting evidence for a role of *PRKAR1B* in neurodevelopmental disorders (NDDs). Statistical analyses of a cumulative dataset of 10,927 cases derived from several NDD patient cohorts found recurrent sites (defined as “*de novo* missense mutations of the same amino acid in two or more unrelated cases”) in *PRKAR1B*, among other potential NDD candidate genes^13^. An analysis of expression profiles and protein-protein interaction^14^ of 253 NDD candidate genes based on the same dataset positioned *PRKAR1B* among a network of genes related to c-Jun N-terminal kinase and mitogen-activated protein kinase cascades, which contained several previously identified NDD candidate genes^15^. Another recent study found a significant enrichment of *de novo* missense variants in *PRKAR1B* in a large sample of 31,058 trio-exomes of children with developmental disorders and their unaffected parents^16^.

*PRKAR1B* is therefore a promising candidate gene for NDDs, including autism spectrum disorder (ASD)^17^, although no clear Mendelian disease association has been established to date. We now report six unrelated individuals with variants of *PRKAR1B*, who share similar features indicative of a novel neurodevelopmental disorder.

## Materials and Methods

### Exome sequencing

Trio-based exome sequencing was performed for individuals #1, #3, #4, #5, and #6. Proband-only based exome sequencing was performed for individual #2, as parental samples were not available. Individuals #1, #2, #3, #5, and #6 were enrolled through GeneDx. Individual #4 was enrolled through the UCLA Clinical Site of the Undiagnosed Diseases Network (UDN). Following informed consent, a comprehensive chart review of medical records was performed.

### Ethics statement

Data collection was performed under the umbrella of human research study H-34578 (Understanding the Molecular Causes of Neuropsychiatric Disease), which was approved by the Baylor College of Medicine Institutional Review Board (IRB). Written informed consent for publication of medical data and images was obtained from the parents or legal guardians of the respective individuals as required by the IRB.

## Results

We report six individuals with variants in the *PRKAR1B* gene (five males, one female; mean age 8.83 years, age range 3–16 as of June 2020). All six individuals were diagnosed with ASD by an expert physician, following DSM-V criteria. Global developmental delay (GDD) was reported in all, and congenital hypotonia was reported in three individuals. All individuals had neurologic anomalies, predominantly disorders of movement. These included dyspraxia/apraxia and clumsiness in all, tremor and dystonia in one, and involuntary movements (eye twitching) in another individual. High pain tolerance was reported by the parents of three individuals, with one individual occasionally harming himself without noticing, and another one sometimes biting himself, when frustrated, to the point of breaking the skin. Only individual #2, whose mother was also reported to have a seizure disorder, manifested seizures.

While speech delay was reported in all individuals, speech regression has been reported in two of them: One individual lost the ability to use two-word phrases, while another one, who previously acquired an active vocabulary of ∼40 words, lost most of it by age two, and became almost nonverbal from the age of five years onwards. Behavioral abnormalities included autistic features like arm/hand flapping, repetitive, and sensory-seeking behavior. Attention deficit hyperactivity disorder (ADHD) was clinically diagnosed in four individuals and suspected by the parents of a fifth. Bouts of aggression were reported in three. Obesity (BMI>30 kg/m^2^) was present in one individual, while another one had a BMI in the borderline obese range (BMI 29.9 kg/m^2^). Obsession with food was also reported in one of the other individuals. A compilation of phenotypic features of each individual can be found in Table 1. No consistent pattern of major malformations or physical anomalies was reported in our small cohort, although one individual had microcephaly and another one had plagiocephaly. The evaluation of facial photographs of individuals #1, #4, #5 and #6 did not reveal any consistent facial dysmorphisms other than upslanting palpebral fissures, which were seen in individuals #1, #2, #4 and #5 (Table 1).

**Table 1:** Genotypes and phenotypic features of individuals #1–6

| Individual # | 1 | 2 | 3 | 4 | 5 | 6 |
| --- | --- | --- | --- | --- | --- | --- |
| **Age range/**<br>**Sex/ BMI** | 10-15y/ M/ Unknown | 15-20y/ F / 29.9 | 5-10y/ M/ Unknown | 5-10y/ M /32.5 | 5-10y/ M/ 17.3 | 0-5y/ M/ 17.9 |
| **Variant**<br>**(NM_001164760)** | c.586G>A, p.Glu196Lys | 500_501delAAinsTT,<br>p.Gln167Leu | c.1003C>T, p.Arg335Trp | c.1003C>T, p.Arg335Trp | c.1003C>T, p.Arg335Trp | c.1003C>T, p.Arg335Trp |
| **De novo?** | Yes | Unknown | Yes | Yes | Yes | Yes |
| **Global**<br>**developmental**<br>**delay** | Yes | Yes | Yes | Yes | Yes | Yes |
| **Diagnosed ASD** | Yes | Yes | Yes | Yes | Yes | Yes |
| **ADHD** | Yes | Unknown | Yes | Suspected by parents,<br>no formal diagnosis | Yes | Yes |
| **Dyspraxia**<br>**/Apraxia** | Yes | Yes | Yes | Yes | Yes | Yes |
| **Congenital**<br>**Hypotonia** | Unknown | Unknown | Unknown | Yes | Yes | Yes |
| **Speech** | Speech delay | Speech delay | Speech delay | Speech regression, non-<br>verbal | Speech delay,<br>1-3 word sentences | Speech delay and<br>regression, single words |
| **Other behavioral**<br>**anomalies** | Aggression | Immature for age | No | Aggression, hand<br>flapping | Arm flapping ,<br>aggression when<br>frustrated | Odd/repetitive<br>behaviors, arm flapping,<br>happy demeanor,<br>sensory-seeking |
| **Pain tolerance** | Normal | Unknown | Unknown | High | Very high | High |
| **Other neurologic**<br>**anomalies** | Tremor, Hemidystonia | Seizures | No | No | No | Eye twitching |
| **Physical**<br>**anomalies** | Upslanting palpebral<br>fissures | Upslanting palpebral<br>fissures, other<br>dysmorphic features<sup>1</sup> | No | Obesity, astigmatism,<br>esotropia, upslanting<br>palpebral fissures, other<br>dysmorphic features<sup>2</sup> | Microcephaly,<br>upslanting palpebral<br>fissures | Torticollis,<br>plagiocephaly,<br>submucosal cleft palate<br>dysmorphic features<sup>3</sup> |
| **Skill regression** | No | Unknown | No | No | No | Potential skill regression<br>and plateauing of<br>progress |
| **Other features** | Unknown | Asthma, fatigue | Sleep problems,<br>nocturnal enuresis | Obstructive sleep<br>apnea, recurrent otitis<br>media | Sleep disturbance,<br>severe eczema* | Restlessness,<br>obstructive sleep apnea |

1 Round face, broad nasal tip, thin upper lip, and short palpebral fissures.
2 Hypotelorism, bitemporal narrowing, epicanthal folds, flat nasal bridge, downturned mouth, tapered fingers, doughy hands, and brachydactyly.
3 Epicanthal folds, slightly posteriorly rotated ears.
\*Individual #5 is also homozygous for a pathogenic variant in *FLG* (c.2282\_2285delCAGT; p.Ser761Cysfs\*36), which explains his severe eczema and dry skin.

*De novo* origin of the respective *PRKAR1B* variants has been confirmed in five individuals, and the parents of individual #2 were not available for testing. Four individuals carry the same variant c.1003C>T (p.Arg335Trp), while the other two carry the missense variants: c.586G>A (p.Glu196Lys) and c.500_501delAAinsTT (p.Gln167Leu) (NM_001164760), respectively. A 3D model of the human R1β subunit structure^18^ is shown in Figure 1B, highlighting the affected amino acid (AA) residues. All reported variants are absent from presumably healthy controls in public databases (gnomAD v2 and v3)^19^. Combined Annotation Dependent Depletion (CADD v1.6) scores^20^ calculated for the single nucleotide variants c.1003C>T and c.586G>A were 26.4 and 24.1, respectively, ranking these variants among the most deleterious 1% of substitutions in the human genome as predicted by the CADD software. The variant c.500_501delAAinsTT is situated close to the 3’ border of exon 5, and predicted functional impacts on splicing vary between the different prediction tools embedded in the Alamut Visual^TM^ variant analysis software (v2.15), ranging from −1.6% (NNSPLICE) to −46.8% (GeneSplicer). To further estimate the functional impact, we calculated Evolutionary Action (EA) scores^21^ of the three variants, which predict functional impacts of AA substitutions by taking into account the phylogenetic distances of AA changes throughout the evolutionary history of the protein, as well as the compatibility of new substitutions with the typical substitutions observed between homologous sequences. The multiple sequence alignment of 106 *PRKAR1B* orthologues and paralogues used in this calculation is shown in Figure S1. The EA scores are normalized on a scale from 0 (predicted wild-type protein activity) to 100 (predicted loss of protein activity), with the value representing the percentage of all possible AA substitutions in the protein that have less impact. The p.Glu196Lys, p.Gln167Leu, and p.Arg335Trp AA substitutions had EA scores of 30, 84, and 87, respectively, indicating strong functional impact for the last two, mostly due to the evolutionary pressure to preserve the Gln167 and Arg335 residues. The relatively low EA score of the p.Glu196Lys substitution is due to higher variation of this AA residue across species (67.6% Glu, 7.6% Asp, 5.7% Pro, 4.8% Ala,4.8% Asn, 2.9% Gln, 1.9% Thr, 1.9% His, 1% Arg and 1% Val), which however still hold a negative or neutral charge, versus the substitution of glutamic acid by positively charged lysine in the affected patient, leading to an actual alteration in charge for the respective residue.

**Figure 1.**
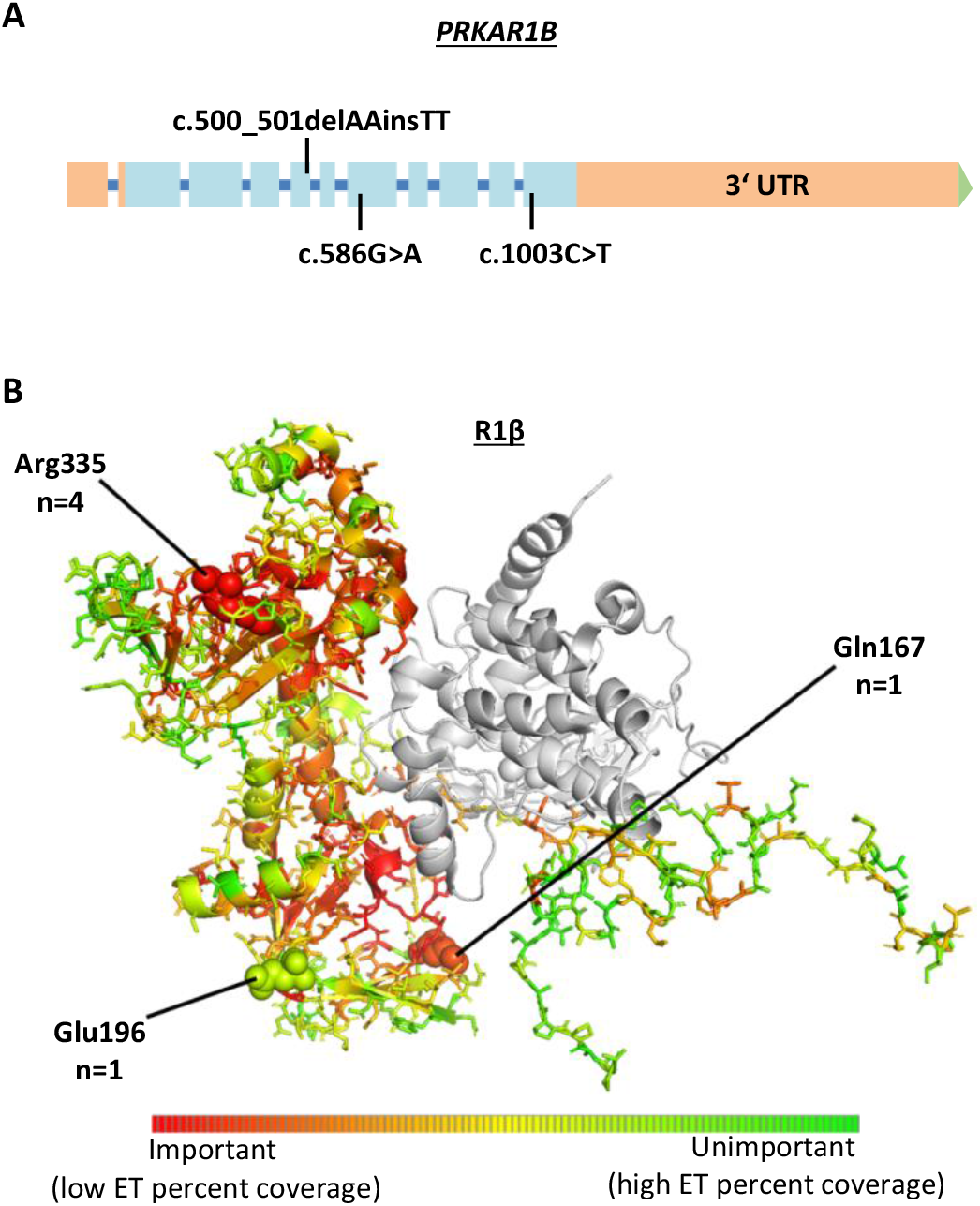
**A**) Distribution of the observed variants within the *PRKAR1B* gene. Exons are shown as boxes, introns as a blue line (introns are not to scale). Light blue color indicates protein-coding sequence **B**) Mutated AA positions within the R1β protein and number of affected individuals. A color shift to red indicates a higher degree of intolerance towards AA variation throughout evolution; according to the respective position’s Evolutionary Trace (ET) score (see Supplemental Methods for further details).

## Discussion

To our knowledge, this study represents the first systematic characterization of a neurodevelopmental disorder associated with missense variants in *PRKAR1B*. In addition to the observed enrichment of *de novo* missense variants of this gene in two large, independent cohorts of individuals with NDDs^15,16^, the recurrent finding of the *de novo* variant c.1003C>T in phenotypically similar individuals strongly suggests that this variant is causative for the observed phenotype (see Supplemental Note 1), and indicates a potential mutational hotspot in the Arg335 residue.

The clinical features of our cohort seem to approximate some aspects of the phenotype of R1β-deficient mice, such as increased pain tolerance^10^, which was reported in three patients carrying the c.1003C>T variant. Furthermore, defects in hippocampal homosynaptic long term depotentiation and low frequency stimulus-induced synaptic depression reported in mice^7^, if also present in humans, may influence synaptic plasticity and thereby potentially impair learning and other cognitive functions. On a cellular level, cognitive abnormalities may be caused by a reduced functionality of the PKA complex and diminished PKA-mediated phosphorylation of CREB in neurons. As the cAMP/PKA/CREB cascade is instrumental for the transcription of memory-associated genes and long-term memory formation^9,22^, further studies might demonstrate functional impacts of *PRKAR1B* variants by measuring CREBP phosphorylation and expression of target genes in human IPSC-derived neurons from affected individuals and healthy controls.

We propose a novel *PRKAR1B*-associated neurodevelopmental disorder with GDD, ASD, neurologic anomalies, and cognitive impairment (no formal IQ scores have yet been obtained from the reported individuals) as principal features. Future research should focus on the functional consequences of *PRKAR1B* variants at a cellular level, as well as the recruitment and clinical characterization of more individuals carrying *de novo PRKAR1B* variants.

## Data Availability

All data supporting the conclusions of our study are included in the manuscript and supplement.

## Acknowledgements

Research reported in this manuscript was partially supported by the NIH Common Fund, through the Office of Strategic Coordination/Office of the NIH Director under Award Number U01HG007703 (SFN, JAM-A, CGSP) and the UCLA California Center for Rare Diseases, within the Institute of Precision Health. The content is solely the responsibility of the authors and does not necessarily represent the official views of the NIH.

The manuscript includes important contributions from On behalf of the Members of the Undiagnosed Diseases Network (see member list on pages 6–8 of the supplement).

## Web resources

The Genome Aggregation Database (gnomAD) https://gnomad.broadinstitute.org/

Combined Annotation Dependent Depletion (CADD) https://cadd.gs.washington.edu/

